# Domestic violence, mental health, and repeat self-harm in Sri Lanka

**DOI:** 10.1101/2021.10.25.21265461

**Authors:** Piumee Bandara, Andrew Page, Thilini Rajapakse, Duleeka Knipe

**Author notes:** **Corresponding author** Piumee Bandara. Authors contributed equally (Thilini Rajapakse, Duleeka Knipe).

## Abstract

**Background:** Evidence on the risk factors for repeat self-harm from low- and middle-income countries is scarce and requires urgent attention.

**Aims:** We sought to examine psychosocial correlates of repeat self-harm in Kandy, Sri Lanka.

**Methods:** Logistic regression models (adjusting for age and sex) were used to examine associations between study factors and repeat self-harm among adult patients (N=292) admitted for self-poisoning.

**Results:** Depression (OR 2.8 95% CI 1.7-4.7), prior psychiatric diagnosis (OR 4.8 95% CI 2.2-10.2), past-year exposure to suicidal behaviour (OR 1.9 95% 1.1-3.3), physical/sexual abuse (OR 3.0 95% CI 1.3-6.9), and psychological abuse (OR 2.3 95% CI 1.3-4.2) were strongly associated with repeat self-harm.

**Conclusions:** Identification and management of domestic violence and psychiatric morbidity should be part of routine clinical assessments of patients presenting with self-harm.

---

Although repeat self-harm is not a strong predictor of future self-harm or suicide at the population level in Asia (Knipe et al., 2019), there may be certain sub-groups who may have an increased risk of suicide following non-fatal self-harm. There remains a paucity of evidence of the risk factors for repeat self-harm from low- and middle-income countries. Psychiatric morbidity has been linked with repeat self-harm in high-income settings (Larkin et al., 2014). Furthermore, patients presenting for self-harm in the UK showed those exposed to domestic violence (DV) were more likely to have a history of self-harm (Dalton et al., 2019). DV has also been previously shown to be strongly associated with self-poisoning in Sri Lanka (Bandara et al., 2020). To our knowledge, no studies have examined the extent to which DV and mental health factors are associated with repeat self-harm in South Asia. Accordingly, we estimated the prevalence of previous self-harm among a sample of Sri Lankan adult patients presenting to hospital for self-poisoning, and examined DV and psychiatric correlates of repeat self-harm.

## Methods

### Data collection

This study is nested within a large case-control study examining childhood adversity and deliberate self-poisoning in the semi-urban town of Kandy, Sri Lanka (Rajapakse et al., 2020). Cases were patients ≥18 years admitted to the medical toxicology ward of a tertiary hospital for deliberate self-poisoning between July 18, 2018 and December 31, 2018. Deliberate self-poisoning (hereafter referred to as self-poisoning) was defined as an act of non-fatal self-poisoning (e.g., pharmaceutical, pesticide, plant) regardless of suicidal intent. Interviews were conducted by trained data collectors in the participant’s preferred language. All research was approved by the appropriate ethics committee (14 June 2018).

### Study variables

Previous self-harm was identified via the question, “Have you ever previously self-harmed in the past?” Depression and harmful alcohol use (i.e., hazardous drinking and/or alcohol use disorder were screened using the Patient Health Questionnaire-9 (PHQ-9) (cut-off score ≥10) and Alcohol Use Disorders Identification Test (AUDIT) (cut-off score ≥8) respectively, both validated for use within the Sri Lankan population (Suraweera et al., 2013). Prior psychiatric diagnosis was ascertained through the question: “Have you ever been diagnosed to have a mental disorder?”; and exposure to suicide/self-harm ascertained through the question: “Did a close family member/close friend you know attempt to harm themselves/ ‘commit’ suicide, during the past one year?” DV was identified using the Humiliation, Afraid, Rape, Kick (HARK) questionnaire and modified to include abuse by any family member living in the household, not just an intimate partner (Bandara et al., 2020). Psychological abuse was categorised as individuals who reported experiencing fear of and/or humiliation by any family member living in the household (without any physical or sexual violence) in the past year, versus no abuse. A composite physical/sexual abuse variable (with or without psychological abuse) was created given the limited number of total sexual violence cases (n=5).

### Statistical analysis

A complete case analysis was conducted (i.e. no missing data). The association between all study factors and repeat self-harm were examined using a series of logistic regression models, adjusting for age and sex. All analyses were conducted in Stata version 16.

## Results

Overall, 298 self-poisoning patients were interviewed. Six participants (2%) had missing data and were excluded, resulting in 292 patients included in the analysis. Overall, 29% (95% CI 24% - 34%) of self-poisoning patients had previously self-harmed. Depression (OR 2.8 95% CI 1.7-4.7), prior psychiatric diagnosis (OR 4.8 95% CI 2.2-10.2), past-year exposure to suicidal behaviour (OR 1.9 95% 1.1-3.3), physical/sexual abuse (OR 3.0 95% CI 1.3-6.9), and psychological abuse (OR 2.3 95% CI 1.3-4.2) were associated with repeat self-harm (Table 1).

**Table 1.** Correlates of repeat self-harm among patients (≥18 years) admitted for self-poisoning, Kandy, Sri Lanka.

|  | History of self-harm |  |  |  |  |
| --- | --- | --- | --- | --- | --- |
|  | Yes (n=84) |  | No (n=208) |  | Odds ratio (95% CI)<sup>a</sup> |
| *Demographic factors* | N | % (95% CI) | N | % (95% CI) |  |
| **Sex** |  |  |  |  |  |
| Male | 41 | 48.8 (37.7-60.0) | 96 | 46.2 (39.2-53.2) | 1.00 |
| Female | 43 | 51.2 (40.0-62.3) | 112 | 53.8 (46.8-60.8) | 0.81 (0.48-1.37) |
| **Age** |  |  |  |  |  |
| 18-25 | 46 | 54.8 (43.5-65.7) | 98 | 47.1 (40.2-54.1) | 1.00 |
| 26-40 | 27 | 32.1 (22.4-43.2) | 67 | 32.2 (25.9-39.0) | 0.85 (0.48-1.50) |
| 41+ years | 11 | 13.1 (6.72-22.2) | 43 | 20.7 (15.4-26.8) | 0.51 (0.24-1.10) |
| **Mental health factors** |  |  |  |  |  |
| **Depression (PHQ-9 ≥10)** |  |  |  |  |  |
| No | 32 | 38.1 (27.7-49.3) | 131 | 63.0 (56.0-69.6) | 1.00 |
| Yes | 52 | 61.9 (50.7-72.3) | 77 | 37.0 (30.5-44.0) | 2.80 (1.65-4.73) |
| **Prior psychiatric diagnosis** |  |  |  |  |  |
| No | 64 | 76.2 (65.7-84.8) | 194 | 93.3 (89.0-96.3) | 1.00 |
| Yes | 20 | 23.8 (15.2-34.3) | 14 | 6.70 (3.7-11.0) | 4.76 (2.23-10.17) |
| **Past-year exposure to suicidal behaviour** |  |  |  |  |  |
| No | 54 | 64.3 (53.1-74.5) | 161 | 77.4 (71.1-82.9) | 1.00 |
| Yes | 30 | 35.7 (25.6-46.9) | 47 | 22.6 (17.1-28.9) | 1.87 (1.07-3.28) |
| **Harmful alcohol use (AUDIT≥8)** |  |  |  |  |  |
| No | 63 | 75.0 (64.4-83.8) | 158 | 76.0 (69.6-81.6) | 1.00 |
| Yes | 21 | 25.0 (16.2-35.6) | 50 | 24.0 (18.4-30.4) | 1.14 (0.55-2.39) |
| **Domestic violence** |  |  |  |  |  |
| **Physical/sexual abuse** |  |  |  |  |  |
| No | 35 | 68.6 (54.1-80.9) | 130 | 83.9 (77.1-89.3) | 1.00 |
| Yes | 16 | 31.4 (19.1-45.9) | 25 | 16.1 (10.7-22.9) | 3.04 (1.34-6.89) |
| **Psychological abuse** |  |  |  |  |  |
| No | 35 | 51.5 (39.0-63.8) | 130 | 71.0 (63.9-77.5) | 1.00 |
| Yes | 33 | 48.5 (36.2-61.0) | 53 | 29.0 (22.5-36.1) | 2.32 (1.30-4.15) |

<sup>a</sup>Odds ratio adjusted for age and sex.
CI=Confidence Interval; PHQ=Patient Health Questionnaire; AUDIT=Alcohol Use Disorder Identification Test.

## Discussion

Exposure to DV, psychiatric morbidity (i.e., depression and a previous psychiatric diagnosis), and suicidal behaviour appear to play a key role in repeat self-harm. The prevalence of repeat self-harm in our study (29%) is higher than higher than what was previously reported in the same hospital roughly 10 years prior (8.7%, 95% CI 6.7–11.0%) (Mohamed et al., 2011). Contrary to the previous study, our study recorded past self-harm regardless of hospital admission. It is likely we captured forms of self-harm (e.g., self-cutting), which would usually not present to hospital. Furthermore, there is increasing evidence in Sri Lanka of a shift in self-harm methods from highly lethal pesticides towards medicinal agents (e.g., paracetamol), which may be less likely to present to hospital (de Silva et al., 2012). Reduced case-fatality may have also resulted in more patients at risk of repetition, highlighting the importance of increasing support for individuals experiencing distress.

The intersections between DV, mental health and suicidal behaviour are complex. A meta-analysis of longitudinal studies reported a bidirectional relationship between DV and suicidal behaviour and similarly for DV and psychiatric disorders (Devries et al., 2013). Qualitative studies from China and Sri Lanka have suggested experience of DV and interpersonal conflict elicits distress and hopelessness, which may translate to self-harm (Marecek & Senadheera, 2012; Wong et al., 2011). The association between exposure to suicidal behaviour and repeat self-harm may reflect shared risk factors, such as exposure to DV, as well as the possible ‘normalisation’ of self-harm as a maladaptive response to stress, but further work is needed to explore this. Alcohol misuse was not associated with repeat self-harm, and one possible reason may have been the greater number of females in the study, who are less likely to misuse alcohol in the Sri Lankan socio-cultural context.

The study is limited by its cross-sectional design, lack of detail regarding the prior self-ham, and potential underestimation of DV. Despite these limitations, given the high rates of DV and the association with psychiatric morbidity in this high-risk group, all patients presenting for self-harm should be assessed for DV and psychiatric morbidity as part of their routine clinical assessment. Further management and support to address potentially co-occurring issues of DV and psychiatric morbidity should be provided as needed. The findings also signal the importance of examination of the impact of exposure to suicidal behaviour. Further qualitative and prospective studies are needed to study the complex intersections between DV, psychiatric morbidity and self-harm.

## Data Availability

Data are available at the University of Bristol data repository, data.bris. Given the sensitivity of the data, only researchers at verifiable institutions will be able to access data.
Any requests will be reviewed by the University of Bristol Access Committee, which includes senior
researchers and representatives from the University. Data will only be released once a controlled data
access agreement has been signed by a nominated institutional signatory.

https://doi.org/10.5523/bris.37pg6mv6x35r12b98aoq4blcgs.

## Notes

### Competing Interest Statement

The authors have declared no competing interest.

### Funding Statement

This work was funded by the UK Medical Research Council (grant number MC_PC_MR/R019622/1) and Wellcome Trust [204813/Z/16/Z]. DK is supported by the Wellcome Trust and Elizabeth Blackwell Institute Bristol.

### Author Declarations

Ethical Review Committee of University of Peradeniya (Faculty of Medicine) gave ethical approval for this work.

## References

Bandara, P., Page, A., Senarathna, L., Kidger, J., Feder, G., Gunnell, D., Knipe, D. (2020). Domestic violence and self-poisoning in Sri Lanka. Psychological medicine, 1–9. https://doi.org/ https://dx.doi.org/10.1017/S0033291720002986

Dalton, T. R., Knipe, D., Feder, G., Williams, S., Gunnell, D., & Moran, P. (2019). Prevalence and correlates of domestic violence among people seeking treatment for self-harm: data from a regional self-harm register. Emergency medicine journal : EMJ, 36(7), 407–409. https://doi.org/ https://dx.doi.org/10.1136/emermed-2018-207561

de Silva, V. A., Senanayake, S., Dias, P., & Hanwella, R. (2012). From pesticides to medicinal drugs: time series analyses of methods of self-harm in Sri Lanka. Bulletin of the World Health Organization, 90, 40–46. https://doi.org/10.2471/BLT.11.091785

Devries, K. M., Mak, J. Y., Bacchus, L. J., Child, J. C., Falder, G., Petzold, M., Watts, C. H. (2013). Intimate partner violence and incident depressive symptoms and suicide attempts: a systematic review of longitudinal studies. PLoS medicine, 10(5), e1001439. https://doi.org/10.1371/journal.pmed.1001439

Knipe, D. W., Metcalfe, C., Hawton, K., Pearson, M., Dawson, A., Jayamanne, S., Gunnell, D. (2019). Risk of suicide and repeat self-harm after hospital attendance for non-fatal self-harm in Sri Lanka: a cohort study. The Lancet Psychiatry, 6(8), 659–666. https://doi.org/10.1016/S2215-0366(19)30214-7

Larkin, C., Di Blasi, Z., & Arensman, E. (2014). Risk factors for repetition of self-harm: a systematic review of prospective hospital-based studies. PLoS one, 9(1), e84282. https://doi.org/ https://doi.org/10.1371/journal.pone.0084282

Marecek, J., & Senadheera, C. (2012). ‘I drank it to put an end to me’: Narrating girls’ suicide and self-harm in Sri Lanka. Contributions to Indian Sociology, 46(1-2), 53-82. https://doi.org/ https://doi.org/10.1177/006996671104600204

Mohamed, F., Perera, A., Wijayaweera, K., Kularatne, K., Jayamanne, S., Eddleston, M., Gunnell, D. (2011). The prevalence of previous self-harm amongst self-poisoning patients in Sri Lanka. Social psychiatry and psychiatric epidemiology, 46(6), 517–520. https://doi.org/ https://doi.org/10.1007/s00127-010-0217-z

Rajapakse, T., Russell, A. E., Kidger, J., Bandara, P., López-López, J. A., Senarathna, L., Knipe, D. (2020). Childhood adversity and self-poisoning: A hospital case control study in Sri Lanka. PLoS one, 15(11), e0242437. https://doi.org/ https://doi.org/10.1371/journal.pone.0242437

Suraweera, C., Hanwella, R., Sivayokan, S., & de Silva, V. (2013). Rating scales validated for Sri Lankan populations. Sri Lanka Journal of Psychiatry, 4(2). https://doi.org/ https://doi.org/10.4038/sljpsyc.v4i2.6320

Wong, S. P., Wang, C., Meng, M., & Phillips, M. R. (2011). Understanding self-harm in victims of intimate partner violence: a qualitative analysis of calls made by victims to a crisis hotline in China. Violence Against Women, 17(4), 532–544. https://doi.org/ https://doi.org/10.1177/1077801211404549

